# Pain management after dental extractions – non-opioid combination analgesics minimize opioid use for acute dental pain

**DOI:** 10.1101/2022.05.12.22274900

**Authors:** Qirong Huang, Linda Rasubala, Richard Gracely, Junad Khan, Eli Eliav, Yanfang Ren

## Abstract

**Objective:** To evaluate long-term changes in pain management strategies and assess the outcomes of opioids and non-opioid combination analgesics after dental extractions.

**Methods:** This is a cross-sectional study of patients who received dental extractions and analgesic prescriptions in a large dental urgent care center in two 12-month periods: January 2012 to December 2012 (Year-2012) and March 2021 to February 2022 (Year-2022). Data extracted from electronic records include type of dental extractions, analgesics prescribed, and follow-up visits. The primary outcome was failure rate measured by the proportions of patients who returned to the clinic for management of pain after receiving dental extractions and analgesic prescriptions.

**Results:** A total of 3,357 patients in Year-2012 and 3,785 patients in Year-2022 received analgesic prescriptions in conjunction with dental extractions. Combination analgesics were significantly higher in Year-2022 (62.5%) than in Year-2012 (34.9%) (RR=1.79, 95% CI 1.70-1.89. p<0.0001). Combinations analgesics were almost exclusively opioids and 1,166 patients, or 34.7%, received opioids in Year-2012, compared to none received opioids, 49.4% received ibuprofen/ acetaminophen and 13.1% received gabapentin combinations in Year-2022. After surgical extractions, a majority were prescribed opioids (52.4%), followed by ibuprofen (46.2%) in Year-2012. In contrast, a majority received ibuprofen/acetaminophen (56.2%) or gabapentin combinations (17.3%) in Year-2022. Ibuprofen/acetaminophen had a failure rate (2.2%) lower than gabapentin combinations (4.4%) (RR=0.50, 95%CI 0.31-0.83. p=0.01), or opioid combinations (21.4%) (RR=0.10, 95%CI 0.08-0.14. p<0.0001). Failure rate for gabapentin combinations was lower than opioids (RR=0.21, 95%CI 0.14-0.31. p<0.0001).

**Conclusions and Relevance:** This study showed a paradigm shift from opioids and single medication analgesics to non-opioids and combination analgesics with ibuprofen, acetaminophen and gabapentin as components in prescribing for pain after dental extractions, which presents an opportunity to minimize or eliminate our reliance on opioids for dental pain.

## Introduction

The epidemic of opioid use disorder (OUD) challenges conventional use of opioids for pain management after dental extractions. Nearly 10 million people reported misuse of prescription opioids in 2019 in the US^1^. Drug-related overdose deaths increased from 70,000 in 2019^2^ to over 100,000 in 2021^3^. Prescription opioids are recognized as the gateway to OUD or misuse of other illicit drugs^4,5^. Dentists frequently prescribe opioids for dental pain^6-8^, which contributes substantially to opioid misuse and OUD^9-11^.

In recognition of the dental role in OUD, the American Dental Association (ADA) recommended the use of prescription drug monitoring programs (PDMP) to promote the appropriate use of opioids, and alternatives such as nonsteroidal anti-inflammatory drugs (NSAIDs) and multimodal strategies for managing postoperative dental pain^12^. Unfortunately, these recommendations have had only a modest effect. Nearly half of the dentists never use the PDMP program^13^ and opioid prescriptions (56.8%) remain higher than non-opioid prescription (43.2%) for acute dental pain^14^.

Prescriptions for pain management after surgical tooth extractions illustrate the current problem: opioid prescription decreased only moderately from 90% in 2011 to about 70% in 2020 in dental clinics affiliated to US dental schools^7,15^, though considerable evidence demonstrates that non-opioid analgesics, such as NSAIDs or a combination of NSAIDs such as ibuprofen with acetaminophen (N-acetyl-para-aminophenol, or APAP), are superior to opioids for dental pain after dental extractions, including third molar surgeries^16-18^. Possible explanations include: 1) immediate-release opioids continue to be the drugs of choice in patients who cannot tolerate NSAIDs or APAP due to direct side effects or indirect effects from interaction with other medications^19^, 2) cases in which NSAIDs are ineffective, 3) dental prescribers are not convinced that short-term opioid prescription may result in opioid dependency^20^, and 4) ingrained prescription habit^21^.

The Howitt Dental Urgent Care (HDUC) at the University of Rochester Medical Center adopted an evidence-based pathway to minimize opioid use for acute dental pain, implemented the PDMP program in 2013, well before the ADA recommendation in 2016, and significantly decreased opioid prescriptions^22^. However, a significant number of patients continued to receive opioid analgesics when NSAIDs or APAP are contraindicated or ineffective.

The OUD epidemic has been exacerbated by an additional public health crisis in the last two years^23,24^. More patients delayed preventive dental care during the COVID-19 pandemic^25,26^, resulting in a significant increase in patients seeking treatments for dental pain. The convergence of the COVID-19 pandemic and the OUD epidemic not only worsens the ongoing crisis related to OUD^27-30^, but also increases difficulties in pain management in dental clinics with rising demand for prescription analgesics when non-opioid alternatives are limited^31^. In response to the clear need for effective non-opioid analgesics, HDUC systematically reviewed existing evidence that inclusion of gabapentin in combination analgesics may benefit patients who could not tolerate either NSAIDs or APAP. Clinical trials have established the efficacy of gabapentin for acute postoperative pain^32-36^, but the evidence on the effectiveness of gabapentin combination analgesics in the real-world dental setting is limited to empirical experiences and clinical impressions.

The purpose of the present study was to compare patterns and effectiveness of analgesic prescriptions for acute dental pain before and after implementation of opioid reduction strategies in HDUC. Effectiveness was evaluated using a real-world measure of proportion of patients returning for additional pain treatment after receiving the prescribed analgesics.

## Methods

This project was approved by the Research Subject Review Board of University of Rochester Medical Center (approval number RSRB-00005798). All patient information were anonymized and de-identified prior to the analysis. This study was a cross-sectional survey of patient records for analgesic prescriptions after dental extractions, and followed the Strengthening the Reporting of Observational Studies in Epidemiology (STROBE) reporting guidelines for cross-sectional studies^37^.

### Data collection

The opioid reduction initiative was first implemented in 2013 with the mandatory requirement of checking the New York State PDMP before prescription of any opioid analgesics in HDUC. A multimodal strategy including gabapentin combination analgesics were initiated in March 2020 during the lockdown period of the COVID-19 pandemic. We used the analgesic prescription data from the past 12 months (March 1, 2021 to February 28, 2022) to represent the current prescription pattern (Year-2022), and the data from the year 2012 (January 1, 2012 to December 31, 2012) to represent the pattern before opioid reduction (Year-2012). Records of all the patients who visited HDUC for tooth extractions and received analgesic prescriptions were analyzed and compared between the two periods.

The following information was retrieved from the electronic records of patients who visited HDUC during the two study periods:

1. Tooth extractions: Extractions were divided into routine or surgical extractions. Routine extraction was defined as removal of erupted tooth or exposed tooth root without raising a soft tissue flap or cutting and removing bone. Surgical extraction was defined as removal of erupted or impacted tooth or buried tooth root that required cutting the soft tissue, and cutting and removing bone structure around the tooth.
2. Classes of analgesics prescribed: Opioid analgesics included hydrocodone, oxycodone, codeine in combination with APAP and/or ibuprofen. Non-opioid analgesics included APAP, NSAIDs (primarily ibuprofen), gabapentin and their combinations. Gabapentin was always used as a combination with either APAP or ibuprofen. Our guidelines for prescribing analgesics have changed with emerging evidence over the years. In Year-2012, patients with mild pain were usually treated with over-the-counter medications including APAP (325mg) and ibuprofen (200mg); those with moderate to severe pain were prescribed higher doses of ibuprofen (400-600mg) or opioid combinations, usually hydrocodone/APAP 5/325mg, and patients expected to have severe pain were prescribed ibuprofen 600-800mg or opioid combinations such as hydrocodone/APAP 7.5/325mg. In Year-2022, patients with mild pain were more likely prescribed with APAP 500mg or ibuprofen 400mg; those with moderate to severe pain were prescribed ibuprofen 600mg or ibuprofen 400mg/APAP 325mg combinations; and those expected to have severe pain were prescribed ibuprofen 400-600mg/APAP 500mg combinations. For patients who had moderate to severe pain but could not take ibuprofen when indicated, they were usually prescribed opioid combinations in Year-2012, but were prescribed gabapentin 300mg/APAP 500mg combinations in Year-2022 when indicated. Similarly, for patients who could not take APAP when combination medications were indicated, hydrocodone/ibuprofen combinations were prescribed in Year-2012, but gabapentin 300mg/ibuprofen 400mg combinations were prescribed in Year-2022.
3. Follow-up visits for postoperative pain: These included patients who had follow-up or observation visits after a procedure performed during a previous visit. Patients who returned to the clinic for additional treatment of pain were identified from the patients with a follow-up observation visit. These patients were considered failed to achieve adequate pain control after receiving tooth extraction and the prescribed analgesics.

### Sample size and statistical analysis

Records of all the patients who visited the HDUC during the two study periods were included in the analyses. Frequencies of dental extractions and different analgesic prescriptions were compared between the two study periods using Chi-square tests. Descriptive statistics, Chi-square tests, relative risk (RR) and 95% confidence intervals (CI) were used to show the change in the pattern of analgesic prescriptions between the two study periods and compare the failure rates of different combination analgesics. All statistics were performed using OpenEpi (Open Source Epidemiologic Statistics for Public Health, Version 3.01) software for two-tailed tests, and a p-value of less than 0.05 was considered statistically significant.

## Results

A total of 8,955 and 14,765 patients visited the HDUC during the two 12-month periods in Year-2012 and Year-2022, respectively. Of these patients, 4,863 (54.3%) received dental extraction in Year-2012, compared to 7,056 (47.8%) in Year-2022 (X^2^=94.68, RR=1.14, 95% CI 1.11-1.17, p<0.0001). A statistically significant lower percentage of patients received dental extractions in the current 12-month periods. The mean age of patients received dental extractions was 37 years (range 18 to 93) in Year-2012, and 39 years (range 18 to 97) in Year-2022. Gender of the patients were about equally distributed between woman and man in both periods (51.6% women and 48.4% man in Year-2012, and 51.0% women and 49.0% men in Year-2022).

In patients who received dental extractions, 3,357 (69.0%) were prescribed analgesic medications in Year-2012, compared to 3,785 (53.6%) in Year-2022 (X^2^=283.9, RR=1.29, 95% CI 1.25-1.32, p<0.0001). A significantly lower percentage of patients received analgesic prescriptions in conjunction with the dental extractions in the current period (Table 1).

**Table 1:** Frequencies of different types of analgesics prescribed for patients received routine or surgical tooth extractions during the two study periods

|  | Year-2012 |  | Year-2022 |  |  |  |
| --- | --- | --- | --- | --- | --- | --- |
| N | 3357 | | 3785 | | $\chi^2$ | p |
| Procedures | n | % | n | % |  |  |
| Routine extraction | 1921 | 57.2 | 2595 | 68.6 | 98.3 | <0.0001 |
| Surgical extraction | 1436 | 42.8 | 1190 | 31.4 |  |  |
| Analgesics |  |  |  |  |  |  |
| APAP | 43 | 1.3 | 1065 | 28.1 | 5732 | <0.0001 |
| Ibuprofen | 2141 | 63.8 | 353 | 9.3 |  |  |
| APAP + ibuprofen | 7 | 0.2 | 1871 | 49.4 |  |  |
| Opioid combinations | 1166 | 34.7 | 0 | 0 |  |  |
| Gabapentin combinations | 0 | 0 | 496 | 13.1 |  |  |

Patterns of analgesic prescription for dental extractions changed drastically between Year-2012 and Year-2022 (Table 1). Overall, combination analgesics were used in a much higher rate in Year-2022 (62.5%) than in Year-2012 (34.9%) (X^2^=541.9, p<0.0001; Rate Ratio=1.79, 95% CI 1.70-1.89). Patients were about 80% more likely to receive combination analgesics than single analgesics after tooth extractions in Year-2022 than in Year-2012. Combination analgesic prescriptions were limited almost exclusively to opioid combinations (99.4%) in Year-2012. A total of 1,166 patients, or 34.7%, received opioid prescriptions after dental extractions in Year-2012. In comparison, none of the patients received opioid prescriptions after extractions in Year-2022, but 49.4% received APAP/ibuprofen combination and 13.1% received gabapentin combinations (Table 1). For patients who were prescribed single analgesics only, ibuprofen was predominant at 63.8% compared to only 1.3% for APAP in Year-2012. This pattern was reversed in Year-2022, when only 9.3% of the patients received ibuprofen compared to 28.1% received APAP (Table 1).

Table 2 and Figure 1 shows the numbers and proportions of different classes of analgesics prescribed for routine extractions and surgical extractions, respectively, in the two study periods. After routine dental extractions, analgesic prescriptions were largely limited to ibuprofen (76.9%) as a single analgesic and opioid combinations (21.5%) in Year-2012, but a much diverse classes of analgesics were used in Year-2022, including APAP (32.1%), ibuprofen (10.4%), APAP/ibuprofen combinations (46.3%) and gabapentin combinations (11.2%). After surgical extractions, a majority of the patients were prescribed opioid combinations (52.4%), followed by ibuprofen as a single analgesic (46.2%) in Year-2012. In contrast, a majority of the patients received APAP/ibuprofen combinations (56.2%), followed by APAP (19.4%), gabapentin combinations (17.3%), and ibuprofen (7.1%) in Year-2022.

**Table 2:** Analgesic prescriptions after dental extractions during the two study periods

|  | Routine Extraction |  | Surgical Extraction |  |
| --- | --- | --- | --- | --- |
| Type of Analgesics | 2012 | 2022 | 2012 | 2022 |
| APAP | 29 | 834 | 14 | 231 |
| Ibuprofen | 1478 | 269 | 663 | 84 |
| APAP/Ibuprofen | 1 | 1202 | 6 | 669 |
| Opioid combinations | 413 | 0 | 753 | 0 |
| Gabapentin combinations | 0 | 290 | 0 | 206 |
| Total | 1921 | 2595 | 1436 | 1190 |

**Figure 1:**
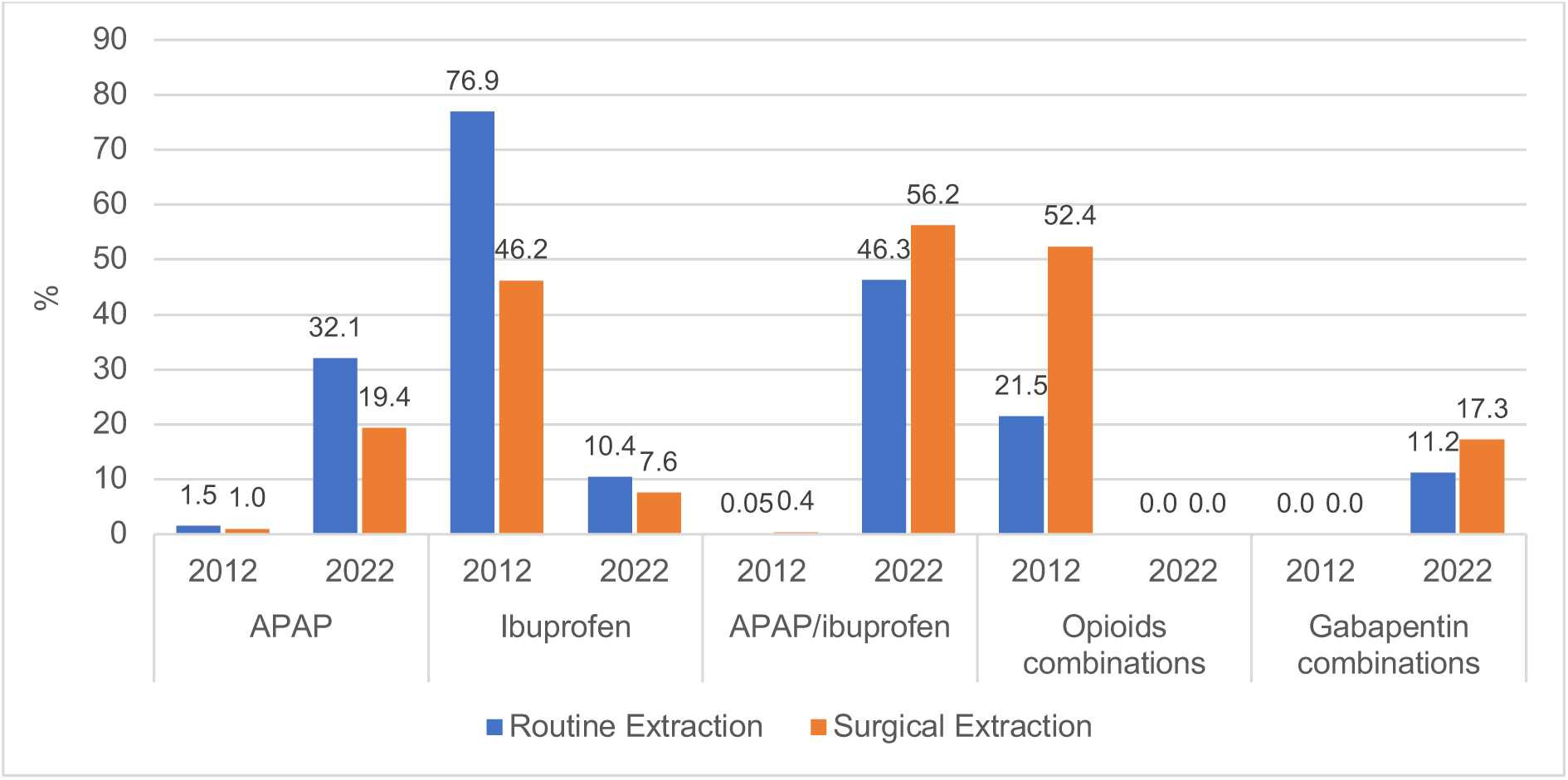
Proportions of different classes of analgesics prescribed (%) after dental extractions (routine and surgical) during the two study periods. Overall, prescription of ibuprofen alone decreased, APAP alone increased, APAP and ibuprofen combinations increased from 2012 to 2022. Opioid combinations were eliminated in 2022 with increased use of APAP/ibuprofen combinations and the addition of gabapentin combinations.

Failure rates for different classes of combination analgesics, defined by the numbers and proportions of patients who returned to the clinic with pain, are listed in Table 3. APAP/ibuprofen combination had a failure rate (2.2%) significantly lower than gabapentin combinations (4.4%) (RR=0.50, 95% CI 0.31–0.83), or opioid combinations(21.4%) (RR=0.10, 95% CI 0.08–0.14). Failure rate for gabapentin combinations was significantly lower than opioid combinations (X^2^=73.1, p<0.0001, RR=0.21, 95% CI 0.14–0.31).

**Table 3:** Pain control outcomes^a^ after simple and surgical extractions in patients received different combination analgesics during the two study periods

|  | Routine Extraction |  | Surgical Extraction |  | Total |  |
| --- | --- | --- | --- | --- | --- | --- |
|  | n | Failure (%) | n | Failure (%) | N | Failure (%) |
| **Non-opioid combinations** | **1493** | **22 (1.5)** | **881** | **42 (4.8)** | **2374** | **64 (2.7)** |
| APAP/ibuprofen | 1203 | 11 (0.9) | 675 | 31 (4.6) | 1878 | 42 (2.2) |
| Gabapentin combinations | 290 | 11 (3.8) | 206 | 11 (5.3) | 496 | 22 (4.4) |
| Gabapentin/ibuprofen | 57 | 3 (5.3) | 44 | 3 (6.8) | 101 | 6 (5.9) |
| Gabapentin/APAP | 233 | 8 (3.4) | 162 | 8 (4.9) | 395 | 16 (4.1) |
| **Opioid combinations** | **413** | **72 (17.4)** | **753** | **177 (23.5)** | **1166** | **249 (21.4)** |
| Codeine/APAP | 98 | 9 (9.2) | 122 | 10 (8.2) | 220 | 19 (8.6) |
| Hydrocodone/APAP | 299 | 58 (19.4) | 619 | 166 (26.8) | 918 | 224 (24.4) |
| Other opioid combinations<sup>b</sup> | 16 | 5 (31.3) | 12 | 1 (8.3) | 28 | 6 (21.4) |

a: Failure" is defined as the number (%) of patients who returned to the clinic with pain after treatment with the prescribed medications. b: Included Hydrocodone/ibuprofen and Oxycodone/APAP

## Discussions

The findings of the present study signify a paradigm shift from opioids and single medication analgesics to non-opioids and combination analgesics in prescribing for pain after dental extractions. Compared to 10 years ago when opioid combinations or ibuprofen alone were the predominant analgesics for dental extractions, a wider selection including APAP, ibuprofen, APAP/ibuprofen combinations and gabapentin combinations are used today to manage postoperative dental pain. This shift represents a more evidence-based approach and presents an opportunity to minimize or even eliminate our reliance on immediate-release opioids for dental pain.

No opioids were prescribed for patients with dental pain in the past 12-months in HDUC. Considering that about 1,800 patients received prescriptions of more than 20,000 pills of opioids annually in this clinic alone before we introduced our opioid reduction initiatives^22^, reducing the reliance on opioids for acute dental pain is particularly salient in light of a persistent OUD crisis^2,38,39^. Population-based cohort studies found that 2 - 7% of the patients receiving a dental opioid prescription would develop new and persist use of opioids within one year^9-11^. A reduction of opioid prescriptions from 1,800 to zero implies that potentially 36 to 126 individuals each year are spared from developing persistent opioid use in this clinic alone.

The reliance on opioid analgesics for dental pain in the US is more likely a cultural phenomenon than a medical necessity. The superiority of non-opioid analgesics such as APAP/ibuprofen combinations to opioids for dental pain has been common knowledge for many years^17,18,22^, yet dentists continue to rely on opioids in North America^10,20,21^. In countries where dentists have limited access to opioid drugs, very few opioids were prescribed for dental pain^40,41^. A 2020 report showed that many patients continued to receive opioids after routine (49.4%) or surgical extractions (70.3%) in an academic dental institution in the US^7^. Compared to the non-opioid alternatives, opioid analgesics also lacked effectiveness, as 65% of the patients who received opioids after routine extraction and 64% after surgical extractions complained of moderate to severe pain in the first week after the procedure, and 17.7% and 11.2% of these patients, respectively, visited or called the clinic due to postoperative pain^7^. The current study corroborated these findings and showed that 21% of the patients received opioid analgesics returned to the clinical for pain, far higher than those received non-opioid combination analgesics. Besides a long list of potential adverse effects, opioids could also aggravate pain by inducing hyperalgesia and increasing sensitivity to pain after both short and long-term use^42-45^. Pharmacogenetic variations related to cytochrome P450 polymorphisms may also render opioids such as hydrocodone or oxycodone ineffective due to poor metabolic conversions^46,47^.

The APAP/NSAIDs combinations, though proven effective, are by no means the silver bullet that will solve the OUD problems associated with prescribing for dental pain. Both NSAIDs and APAP have inherent shortcomings because of their systematic toxicities on vital organs and physiological functions^48,49^. Randomized controlled trials that demonstrated efficacies of APAP, NSAIDs and their combinations on dental pain used the third molar surgery model for postoperative pain in young and healthy volunteers^17,18^. As clinicians facing patients of all ages and health status, we treat many patients who have contraindications to NSAIDs or APAP, either due to compromised systematic health or potential drug-drug interactions with their existing medications^50^. For patients who have contraindications to either NSAIDs or APAP and those who could not achieve adequate pain relief from the APAP/NSAIDs combination, opioids become the obvious and only choice because there are no other non-opioid alternatives for acute dental pain^19^.

Gabapentinoids, including gabapentin and pregabalin, are structural derivatives of γ amino butyric acid and act on both central and peripheral sensory pathways to inhibit central sensitization and peripheral pain transmission^51^. Though gabapentinoids are classified as antiepileptics and were approved primarily for neuropathic pain such as postherpetic neuralgia and fibromyalgia, numerous clinical trials have demonstrated their efficacies for acute pain and for reduction of opioid consumption after major surgery^32,33,52^. A Cochrane review demonstrated efficacy of gabapentin for acute pain after dental extractions^32^. A recent systematic review indicates that preoperative use of gabapentinoids reduces pain and opioid consumption after various oral surgeries including third molar removals^33^. Gabapentioids are not metabolized in the body and is excreted through the kidney in its original form, providing safety for combination with other analgesics^53^. Such combinations may provide additive or synergistic analgesic effects^35,54,55^. Gabapentin as a single medication is probably not adequate for acute dental pain as it has a number-needed-to-treat (NNT) value of 11 for dental pain^32^, but it compares favorably against codeine, a common opioid analgesic, which has an NNT of 21 for dental pain^56^. Codeine in combination with APAP is frequently used for dental pain^10^. Gabapentin used in combination with APAP or NSAIDs should provide a potential alternative to opioids^35,54,55^, especially when the APAP/NSAIDs combination is contraindicated or ineffective.

Pregabalin has the same mechanisms of actions with gabapentin but may have advantages due to its favorable pharmacodynamic properties^57^. We elected to use gabapentin because its efficacy against dental pain is established in randomized controlled trials^32^. The data from the present study provides additional evidence that gabapentin when used in combination with APAP or ibuprofen may be potentially effective against acute dental pain. However, this data must be treated with caution because of its retrospective nature. The outcome measure used in this study, the number of patients returned to the clinic for treatment of pain after receiving dental extractions and the analgesics, is too crude to arrive at definitive conclusions of effectiveness. Well-designed clinical trials are needed to assess the effectiveness and safety in patients with a wide range of age and health status. Gabapentin is not FDA-approved for acute pain and it may have significant adverse effects^58,59^. Considering that gabapentin is one of the most prescribed medications in the US and up to 95% of its prescriptions were “off-label”^60,61^, its use should be judicious and based on thorough understanding of the best available evidence. Patients should be informed the off-label status and potential adverse effects. In addition, gabapentin may potentiate the effects of opioids and have its own potential for misuse or abuse^62^. We have limited its use to a maximum of 600mg per day divided in 2 doses for 3 days, which is very small in quantity and duration compared to greater than 1200mg per day for 8 weeks that are often used for chronic pain conditions^59^. We believe that the potential benefits of its short-term use outweigh its potential risks in comparison to opioids for patients with moderate to severe postoperative dental pain.

The COVID-19 pandemic has significantly affected oral health and changed dental care to a more palliative approach worldwide^25,31^. A much larger volume of patients visited HDUC but a lower percentage received dental extractions compared with previous years. The fact that less patients received prescription analgesics after dental extractions in the current period also reflected the effect of the pandemic on dental care – more patients had already received analgesics elsewhere before they arrived at our clinic for dental extractions. We do not deny that opioid analgesics are effective for many patients and have their rightful roles as part of our armamentarium against pain. Our efforts in minimizing opioid use for dental pain is based on our assessments of potential risks and benefits and our professional judgement. Safe and effective pain relief remain our primary goal for patients with dental pain.

In summary, gabapentin combinations with APAP or ibuprofen may provide viable alternatives to opioids for acute dental pain. But its effectiveness and safety awaits confirmation from well-designed clinical trials. We do not advocate wide-spread use of gabapentin combinations for acute dental pain before more definitive evidence is available, though we believe that their judicious use may benefit a select group of patients.

## Data Availability

All data produced in the present work are contained in the manuscript

